# Functional annotation with expression validation identifies novel metastasis-relevant genes from post-GWAS risk loci in sporadic colorectal carcinomas

**DOI:** 10.1101/2023.06.13.23291271

**Authors:** Lai Fun Thean, Michelle Wong, Michelle Lo, Iain Tan, Evelyn Wong, Fei Gao, Emile Tan, Choong Leong Tang, Peh Yean Cheah

## Abstract

Colorectal cancer (CRC) is the third highest incidence cancer and leading cause of cancer mortality worldwide. Metastasis to distal organ is the major cause of cancer mortality. However, the underlying genetic factors are unclear. This study aims to identify metastasis-relevant genes and pathways for better management of metastasis-prone patients. Multiple lines of evidence have indicated that germline variants play important role in shaping the somatic (tumor) genome. A case-case genome-wide association study comprising 2677 sporadic Chinese CRC cases (1282 metastasis-positive vs 1395 metastasis-negative) was performed using the Human SNP6 microarray platform and analyzed with the correlation/trend test based on the additive model. Single nucleotide polymorphism (SNP) variants with association testing -log10p-value ≥ 5 were imported into Functional Mapping and Annotation (FUMA) for functional annotation which uncovered glycolysis as the top hallmark geneset. Transcripts from two of the five genes profiled, HAX1 and HMMR, were significantly down-regulated in the metastasis-positive tumors. In contrast to disease-risk variants with minimal impact on survival, HAX1 appeared to act synergistically with HMMR in significantly impacting metastasis-free survival. Furthermore, examining the subtype datasets with FUMA and Ingenuity Pathway identified distinct pathways demonstrating sexual dimorphism in CRC metastasis. Combining genome-wide association testing with in silico functional annotation and wet-bench validation identified metastasis-relevant genes that could serve as features to develop subtype-specific metastasis-risk signatures for tailored management of Stage I-III CRC patients.

## 1. Introduction

Colorectal cancer (CRC) is the third highest incidence cancer and leading cause of cancer mortality worldwide [1]. In Singapore, it is the most frequent cancer for men and women combined and the second leading cause of cancer-related death (https://www.nrdo.gov.sg/). Like most cancers, metastasis to distal organ such as liver or lung is the major cause of CRC mortality. This trend is expected to be exacerbated by an aging population in most developed countries as aging is a major risk factor for cancer [2]. Although early-stage (Stage I-II) CRCs with tumors confined to the colonic walls are supposed to be curative after surgery, up to 25% eventually succumb to metastasis [3]. About 50% of lymph-node involved Stage III CRCs succumb to metastasis, usually within 5 years from surgery. Stage IV CRCs are metastasis-positive cases with distal organ involvement at diagnosis.

Most CRC (80%) is sporadic, occurring in individuals aged 50 or more, where environmental factors such as diet and exposure to carcinogens and genetic susceptibility would contribute to disease occurrence [4-6]. Similarly, genetic predisposition is likely to play important role in disease progression and metastasis. Nevertheless, the under-lying germline genetic factors for metastasis are currently unclear.

We aim to identify germline metastasis risk variants in sporadic CRC by case-case genome-wide association study (case-case GWAS). For the past few decades, researchers have focused on case-control GWAS and successfully identified scores of common single nucleotide polymorphism (SNP) risk variants for CRC occurrence by comparing the genomes of patients to that of healthy controls (https://www.ebi.ac.uk/gwas/) [7]. In contrast, case-case GWAS study allows comparison of germline variants between metastasis-positive and metastasis-negative cases. The major limiting factor for such an approach is the ability to accrue cases with definitive metastasis status.

We successfully identified genomic metastasis risk loci that were distinct from the genetic susceptibility loci for disease occurrence. Moreover, these metastasis risk variants have higher effect sizes (odds ratio, OR>1.2) suggesting that germline risk variants could play greater role for disease progression than disease occurrence. Functional annotation on the overall dataset identified glycolysis pathway as the top hallmark geneset. Differential expression and association with metastasis-free survival validated two of the five genes implicated in the glycolysis geneset. Further analysis enabled discovery of metastasis-relevant genes and distinct pathways for different disease subtypes. Validated metastasis-relevant genes can serve as features for the development of subtype-specific metastasis risk signatures with high positive predictive values to further stratify Stage I-III CRC patients for better management of metastasis-prone patients [8].

## 2. Materials and Methods

### 2.1. Clinical samples

A sporadic CRC case was defined as a patient aged 50 years or more at date of operation and without dominant family history of familial adenomatous polyposis or Lynch Syndrome. A total of 2,500 morphologically normal mucosa specimens and 500 blood samples of sporadic CRC cases of Chinese descent from Singapore General Hospital (SGH) and National Cancer Center Singapore (NCCS) respectively were selected as cases for the genome-wide discovery panel. Both SGH and NCCS are located on the same SingHealth Campus. Patients usually have colorectal surgery at SGH and are followed up by medical oncologists at NCCS if necessary. Matched mucosa specimens were sampled ≥ 5 cm from the tumors within 30 min of surgery, snap-frozen and archived in the Department of Colorectal Surgery Tissue Repository, SGH. Samples included were collected from 1996 - 2018. In addition, another 837 cases were recruited as the independent replication panel (RP) from the two centers. About 10% of the RP cases were collected from NCCS.

All cases were defined as metastasis-positive or metastasis-negative using clinically documented data. Metastasis-positive status of the patients is confirmed based on distal organ involvement attributable to primary CRC, either from histopathological report or computed tomography/positron emission tomography (CT/PET) scan regard-less of tumor staging at the time of operation. Metastasis-negative status is confirmed with at least 5 years of follow-up when there is no distal organ involvement. Cases whose metastasis status cannot be confirmed definitively were excluded from analysis. We also interrogated the cause of death with the National Registry of Diseases Office. Cases where cause of death due to cancer cannot be confirmed were excluded.

Other clinical pathological features such as gender, tumor stage, site and distal organ site were similarly extracted from the clinical database (Table 1). Left-sided and right-sided CRC tumors are defined as tumors to the left and right of the splenic flexure respectively.

**Table 1.**
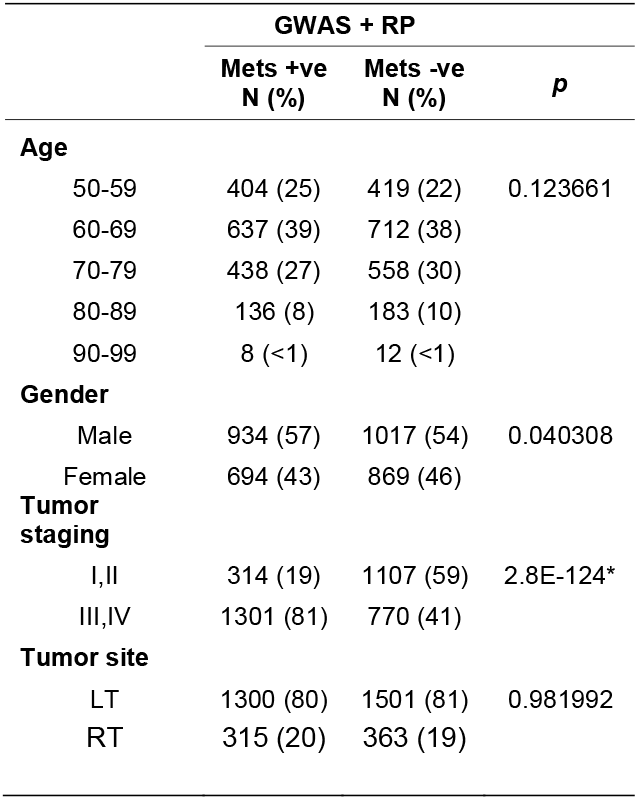
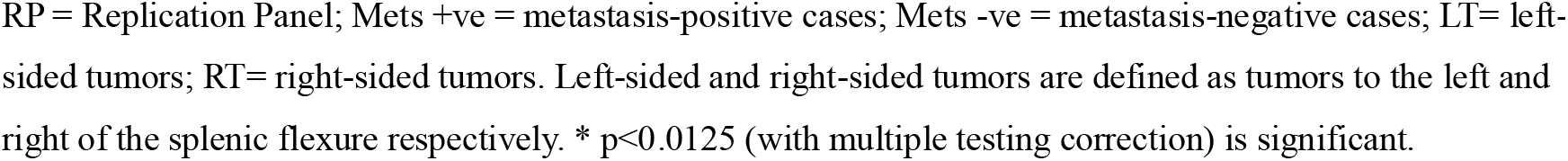
Clinicopathological Features of metastasis-positive and metastasis-negative patients in the pooled GWAS and Replication panels.

|  | GWAS + RP |  | *p* |
| --- | --- | --- | --- |
|  | Mets +ve<br>N (%) | Mets -ve<br>N (%) |  |
| **Age** |  |  |  |
| 50-59 | 404 (25) | 419 (22) | 0.123661 |
| 60-69 | 637 (39) | 712 (38) |  |
| 70-79 | 438 (27) | 558 (30) |  |
| 80-89 | 136 (8) | 183 (10) |  |
| 90-99 | 8 (<1) | 12 (<1) |  |
| **Gender** |  |  |  |
| Male | 934 (57) | 1017 (54) | 0.040308 |
| Female | 694 (43) | 869 (46) |  |
| **Tumor staging** |  |  |  |
| I,II | 314 (19) | 1107 (59) | 2.8E-124* |
| III,IV | 1301 (81) | 770 (41) |  |
| **Tumor site** |  |  |  |
| LT | 1300 (80) | 1501 (81) | 0.981992 |
| RT | 315 (20) | 363 (19) |  |

RP = Replication Panel; Mets +ve = metastasis-positive cases; Mets -ve = metastasis-negative cases; LT= left-sided tumors; RT= right-sided tumors. Left-sided and right-sided tumors are defined as tumors to the left and right of the splenic flexure respectively. \* $p < 0.0125$ (with multiple testing correction) is significant.

### 2.2. Genome-wide scan, quality assurance filtering and genotype calls

Genomic DNA was extracted using the Qiagen DNeasy blood and tissue kit according to the manufacturer’s protocol. Whole genome scan was performed with 600ng of DNA using AffymetrixTM Genome-Wide Human SNP Array 6.0 featuring 906,000 SNPs and 946,000 copy number probes. Genotype calls and quality controls including the Contract Quality Control (CQC) metric were performed using the Genotyping Console 4.0 software. CQC analysis was performed for each batch of samples and mean of the CQC was calculated for samples that passed the 0.4 threshold. Samples with a CQC <0.4 were either repeated or excluded. We performed statistical assessments of experimental qualities for the SNP6 array and make genotype calls with the Analysis Power Tools (APT) program on 2,833 samples. SNP quality check was performed with three components of SNPolisher under APT: ps-metrics, ps-classification and OTV (off target variants) caller. SNPs that passed all the QC requirements were 841,144 SNPs. The apt-package-util and apt-probeset-genotype were performed to create 2,738 CHP files for further analysis. Details of the scan, quality filtering and genotype calling are in the online supplementary methods (Supplemental File S1).

### 2.3. Whole genome association analysis

The CHP files were imported into Golden Helix SVS for further quality filtering and association testing. Forty-three samples with low call rate (autosomes) of < 0.90, inconsistent gender and cryptic relatedness were excluded. Principal Component Analysis (PCA) was performed with 270 HapMap samples and 268 individuals from Chinese, Malay, and Indian population from the Singapore Genome Variation Project (SGVP) to assess for population stratification. PCA was also performed to correct for batch effects and the pattern of variation across samples. Further SNP quality assurance was performed to exclude SNPs with call rate < 0.9, minor allele frequency < 0.01, number of alleles > 2. The total number of SNPs included for the association tests were 647,545 SNPs. Data output was used for Quantile-quantile (Q-Q) plot. Genomic Control was performed to measure the inflation factor, λ, based on the chi-square statistics. Eighteen more samples were excluded due to unconfirmed clinicopathological features. Genotype association testing was performed on 2,677 filtered samples using the additive model as the genetic model and Correlation/Trend test as the statistical method. Association testing was independently performed on disease subtype datasets using the same statistical method.

### 2.4. Functional annotation and pathway analysis

FUMA is an integrative platform using information from 18 different biological resources including regional linkage disequilibrium patterns, expression quantitative trait loci (eQTLs) and chromatin information to facilitate functional annotation of GWAS results, gene prioritization and interactive visualization [9]. Genetic variants not on the microarray platform were imputed using East Asians as reference. The GWAS SNP summary statistics were imported into FUMA SNP2GENE workflow to define the genomic risk loci and annotate the candidate SNPs. These SNPs were mapped to genes by ANNOVAR annotation. Details of the input parameters for the SNP2GENE workflow are in the online supplementary methods (Supplemental File S1). Mapped genes were submitted into GENE2FUNC for gene set enrichment analysis (GSEA). Enrichment of prioritized genes in biological pathways was tested against genes reported in Molecular Signatures Database (MsigDB) v7.1 and WikiPathways using the hypergeometric test. P value (P < 0.05) was corrected with the Benjamin-Hochberg method.

The mapped gene sets for the overall and disease subtypes were also uploaded into Ingenuity Pathway Analysis (IPA) software for canonical pathways and comparison analyses [10]. Canonical Pathways were identified by Fisher’s exact test and clustered using a hierarchical agglomerative clustering algorithm based on average linkage and Euclidean distance metric.

### 2.5. Multiplex SNP genotyping assay

SNP Genotyping was performed on 23 selected top SNPs for the RP with Fluidigm 192.24 Dynamic Array Integrated Fluidic Circuit (IFC). Selected SNPs were uploaded to Fluidigm D3 web-based assay design tool to customize SNP Type assays, based on allele-specific PCR SNP detection with a universal probe set. Specific target amplification (STA) was conducted on all samples for DNA enrichment before loading onto the IFC array. IFC array was injected with amplified DNAs and SNP Type assays. Priming and loading was run on IFC Controller RX and real-time PCR was performed on BioMark HD system with touch down PCR from 64-61°C, dropping 1°C per cycle followed by additional 34 PCR cycles at 60°C. All the PCR conditions are based on the manufacturer’s protocol pre-set under SNPtype 192×24 v1 program.

### 2.6. qRT-PCR expression analysis

Tumor samples were routinely enriched for tumor cells (≥75%) before RNA extraction. RNA was extracted from homogenized fresh frozen tissues in liquid nitrogen using the Qiagen RNeasy kit according to manufacturer’s protocol. Duplex real time qPCR (for gene of interest and ACTB as endogenous control) was performed on 188 Stage III samples from the GWAS panel using C1000 thermal cycle with CFX384 real time (4-color) system (Bio-Rad, Hercules, CA) on 384 well plates. The PrimePCR™ Probe and Control Assay ID and PCR conditions were detailed in the online supplementary methods (Supplemental File S1). The CFX Maestro Software 1.1 was used for data collection and analysis. Relative expression in the matched mucosa and tumors compared to endogenous control ACTB was calculated using the comparative Ct (ΔCt) method. ΔCt = Ct_goi_ – Ct _ACTB_.

### 2.7. Statistical Analysis

Statistical tests were performed using Stata (College Station, Texas, USA). Two sample t-test for unequal variance was carried out to compare gene expression levels between patients with or without metastasis.

Survival time to metastasis is time from surgery to first clinical documentation of metastasis or if metastasis-free, to December 31, 2018. The metastasis free survival was plotted using Kaplan-Meier method and the survival difference between gene expressions was tested using Log-rank test. Chi-square test statistics were used to compare the clinicopathological features of metastasis-positive and metastasis-negative cases (Table 1). Logistic regression was performed to compute the risk of metastasis-positive cases compared to metastasis-negative cases in pooled GWAS and RP panels (Table 2).

**Table 2.** Metastasis risk variants with increased significance in pooled GWAS and RP.

| Chr | Associated genes | rsID | Analyses | GWAS |  |  |  | GWAS + RP |  |  |  |  |
| --- | --- | --- | --- | --- | --- | --- | --- | --- | --- | --- | --- | --- |
| | | | | N | Met+* | Met-** | Corr/Trend $-\log_{10} P$ | N | Met+* | Met-** | Corr/Trend $-\log_{10} P$ | OR (95% C. I.)^ |
| 5q34 | Upstream CCNG1 HMMR MAT2B | rs7711080 | I/II/III/IV# | 2677 | 1282 | 1395 | 5.97 | 3514 | 1628 | 1886 | 6.79 | 1.45 (1.24, 1.71) |
| 1q21.3 | ADAR | rs2131902 | I/II/III/IV | 2677 | 1282 | 1395 | 5.08 | 3514 | 1628 | 1886 | 6.43 | 0.81 (0.70, 0.93) |
|  |  |  | Liver+/lung mets | 2480 | 1085 | 1395 | 6.10 | 3224 | 1359 | 1886 | 6.86 | 0.82 (0.71, 0.95) |
| 8q13.3 | KCNB2 | rs2919408 | Bone mets (Left-sided) | 1257 | 133 | 1124 | 6.59 | 1647 | 146 | 1501 | 7.43 | 1.99 (1.35, 2.93) |

## 3. Results

### 3.1. Characterization of GWAS Discovery and Replication Panels

There were no significant differences in the age distribution, gender and tumor sites of metastasis-positive and metastasis-negative cases in the pooled GWAS and RP panels (Table 1). As expected, tumor stages which serve as the surrogate endpoint for metastasis is very significantly different between metastasis-positive and metastasis-negative cases (p=2.8E-124). Treatment follows the standard protocol for all patients regardless of metastasis status. At the time of sample collection within an hour post-surgery, the metastasis status of stage I-III cases would not have been known. Most (80%) of the CRC cases were aged 55-79 years. The major (2/3) metastatic sites were liver and lung, followed by peritoneal and bone metastasis which constituted about another 20%. There were also no significant differences in the frequencies of the cases stratified by age group, gender, tumor stage, tumor site, metastatic status, and site between the GWAS and RP panels. The gender, tumor stage and site distribution of the cases recruited for both panels were representative of the local CRC incidences.

### 3.2. Genome-wide scan, quality assurance filtering

The GWAS CHP files that passed the Affymetrix quality filtering were imported into Golden Helix SVS and subjected to further quality assurance filtering (overall genotypic call rate of >0.9, minor allelic frequency of > 1%) and Principal component analysis (PCA). PC plot with the Singapore Genome Variant Project (SGVP) and Hap-Map subjects comprising various ethnicities indicates that there is no population sub-structure (Supplemental Figure S1A). All our cases clustered with the Chinese subjects in the HapMap as well as the SGVP. The PC plots indicate both metastasis-positive and metastasis-negative as well as male and female cases are evenly distributed (Supple-mental Figure S1B and S1C). The Q-Q plot (y=x) also indicates no significant evidence of allelic test statistic inflation i.e., good genomic control (λ=1.003) without the need for any PC correction (Supplemental Figure S1D).

### 3.3. Overall and subtype association testing

We performed genome-wide SNP association testing on the final dataset of 2677 cases (1282 metastasis-positive vs 1395 metastasis-negative cases) using the additive model and discovered several regions of interest (ROI) with the top candidate locus at chromosome 20p12 with –log10p-value of 6.13 (p=7.41E-07), followed by ROIs at chromosomes 5q34, 2p13.3, 4q12, and 1q21 that have –log10p-values of >5.0 (p≤1E-05) (Figure 1A, Supplemental Table S1).

**Figure 1.**
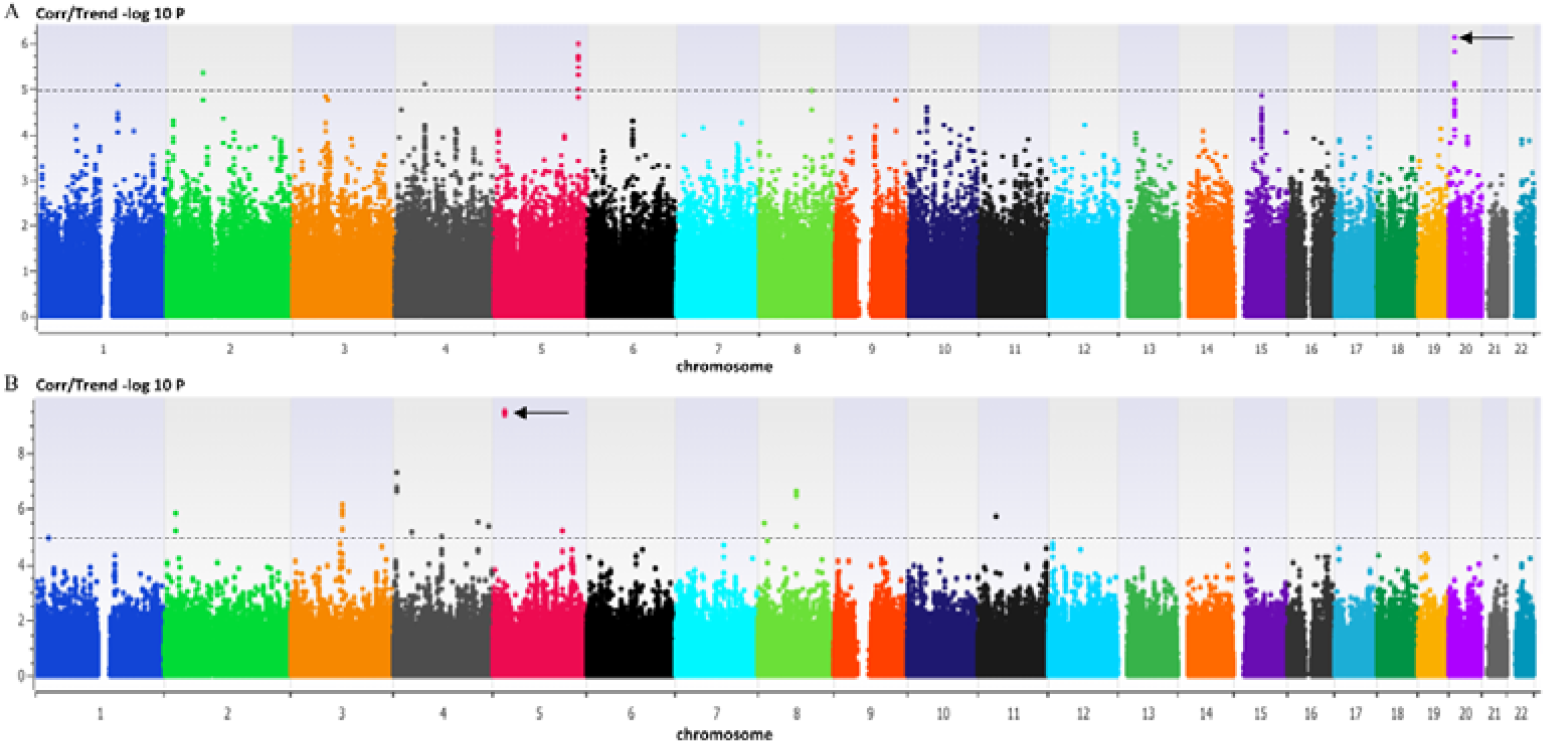
Manhattan Plot of overall vs subtype analysis. A) Overall metastasis-positive (n = 1282) vs metastasis-negative (n = 1395); B) Bone-metastasis (n=150) vs metastasis-negative cases (n=1395). Arrow indicates top SNP with highest –log10p-value for the association testing.

We created subtype-specific datasets by gender, tumor stage, tumor site and distal organ site and independently performed SNP association testing on these datasets. We discovered that the –log10 p-value of association testing of the overall dataset is not always higher than the various subtypes with lower sample sizes suggesting that the variants could be specific to the subtype. For instance, when metastasis to other distal organs (such as liver, lung, peritoneum) were omitted in the association testing of bone-metastasis vs metastasis negative cases, the –log10 p-value for rs16889360 increased from 2.06 to 7.97 despite a much-reduced sample size (Figure 1B, Supplemental Table S2). This indicates that the rs16889360 is a bone-specific risk variants and inclusion of metastasis to other distal organs in the metastasis-positive cases serve as ‘contaminants’ in the associating testing.

### 3.4. Functional annotation using FUMA

We imported candidate risk SNPs with summarized statistics -log10 p ≥ 5 (p≤1E-05) into Functional Mapping and Annotation (FUMA) software for ‘SNP2GENE’ and ‘GENE2FUNC’ analysis (Supplemental Figure S2).

Using FUMA ‘SNP2GENE’ function, we identified 5 genomic risk loci and 101 mapped genes based on gene set enrichment analysis and the Molecular Signatures Database for the overall GWAS dataset (Supplemental Figure S3A-C). 74 of these are protein-coding genes. The histogram displays the proportion of SNPs which have corresponding functional annotation assigned by ANNOVAR relative to all SNPs in the East Asian reference panel (Supplemental Figure S3B). Majority (>60%) of the SNPs are in the intronic regions of genes. The distribution of the SNPs and mapped genes in the 5 genomic risk loci are represented in Supplemental Figure S3C.

Various genesets or pathways were associated with these mapped genes by FUMA ‘GENE2FUNC’ analysis. These genesets were implicated in different metastasis steps, from epithelial-mesenchymal transition (EMT) to mechanical stress in circulation to implantation at distal organs. The top hallmark geneset identified in the ‘GENE2FUNC’ workflow is glycolysis, with 5 genes in four different genomic loci (Supplemental Figure S3D).

We expression-profiled these 5 genes in 188 (94 metastasis-positive vs 94 metastasis-negative) cases from the GWAS panel by quantitative real-time RT-PCR assay. The mean ΔCt values of hyaluronan-mediated motility receptor (HMMR, also known as RHAMM) and hematopoietic substrate 1 associated protein X 1 (HAX1) were significantly higher in the metastasis-positive tumors (indicating lower expression) compared to the metastasis-negative tumors (Figure 2A). Kaplan-Meier survival analysis as a function of the median ΔCt value of the two genes were performed. These plots indicate that lower expression of both genes in the metastasis-positive tumors was significantly correlated with worse metastasis-free survival (Figure 2B and 2C).

**Figure 2.**
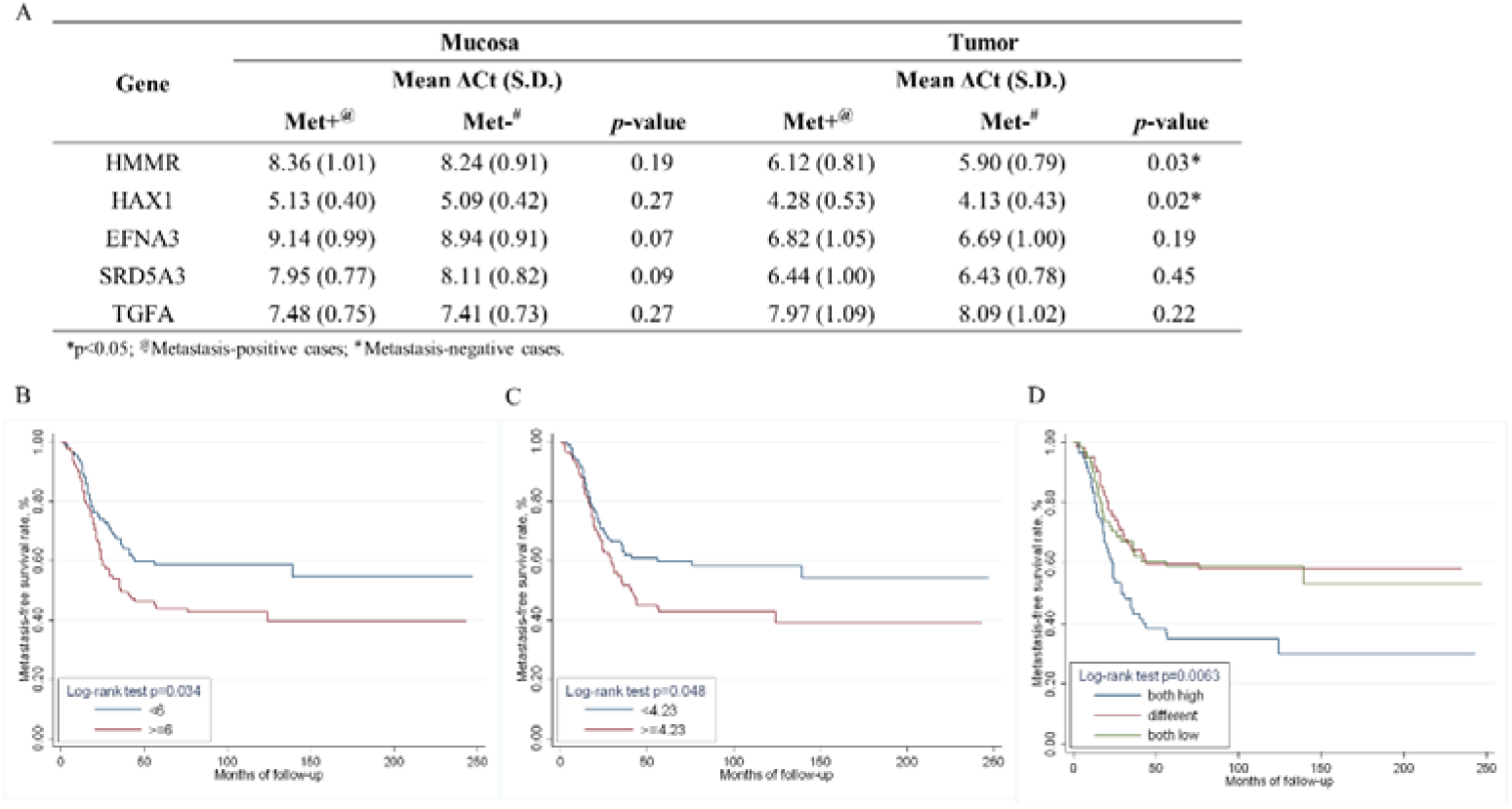
HMMR and HAX1 transcript expressions in the tumors are correlated with metastasis status. A) Two sample t-test for expression of the 5 glycolytic genes in metastasis +ve vs metastasis –ve cases. (B-D) Kaplan-Meier plots for HMMR and HAX1 with Metastasis-free survival (Y-axis). Cut off is by the median ΔCt value. B) HMMR High (>=6) or Low (<6) ΔCt; C) HAX1 High (>=4.23) or Low (<4.23) ΔCt; D) Both genes with high ΔCt; either is high (different) or both genes are low ΔCt. High ΔCt implies low expression of the gene.

Moreover, the expression of the two genes appear to act synergistically in promoting metastasis in CRC as cases with both genes at low expression have significantly lower metastasis-free survival than cases with either one or both genes at low expression (Figure 2D).

We interrogated further the disease subtype datasets with FUMA, focusing on the Gene Ontology categories of biological processes, and molecular functions for gender-, tumor stage- and distal-organ specific metastasis. Surprisingly, many different genesets ranging from immune response to epidermal cell differentiation were enriched in the female but not male patients (Figure 3A). The enriched genesets for left-sided early stage (I/II) CRCs focused on TGF-β/HOX mediated functions while the only function highlighted for left-sided stage III CRCs is ephrin receptor binding (Figure 3B). The enriched pathways for left-sided liver and/or lung-specific metastasis were the triglyceride lipase and carboxylic ester hydrolase functions, similar to those of left-sided stage I-IV CRCs (Figure 3B) while keratinization and negative regulation of muscle relaxation were most enriched for peritoneal and bone metastasis respectively (Figure 3C).

**Figure 3.**
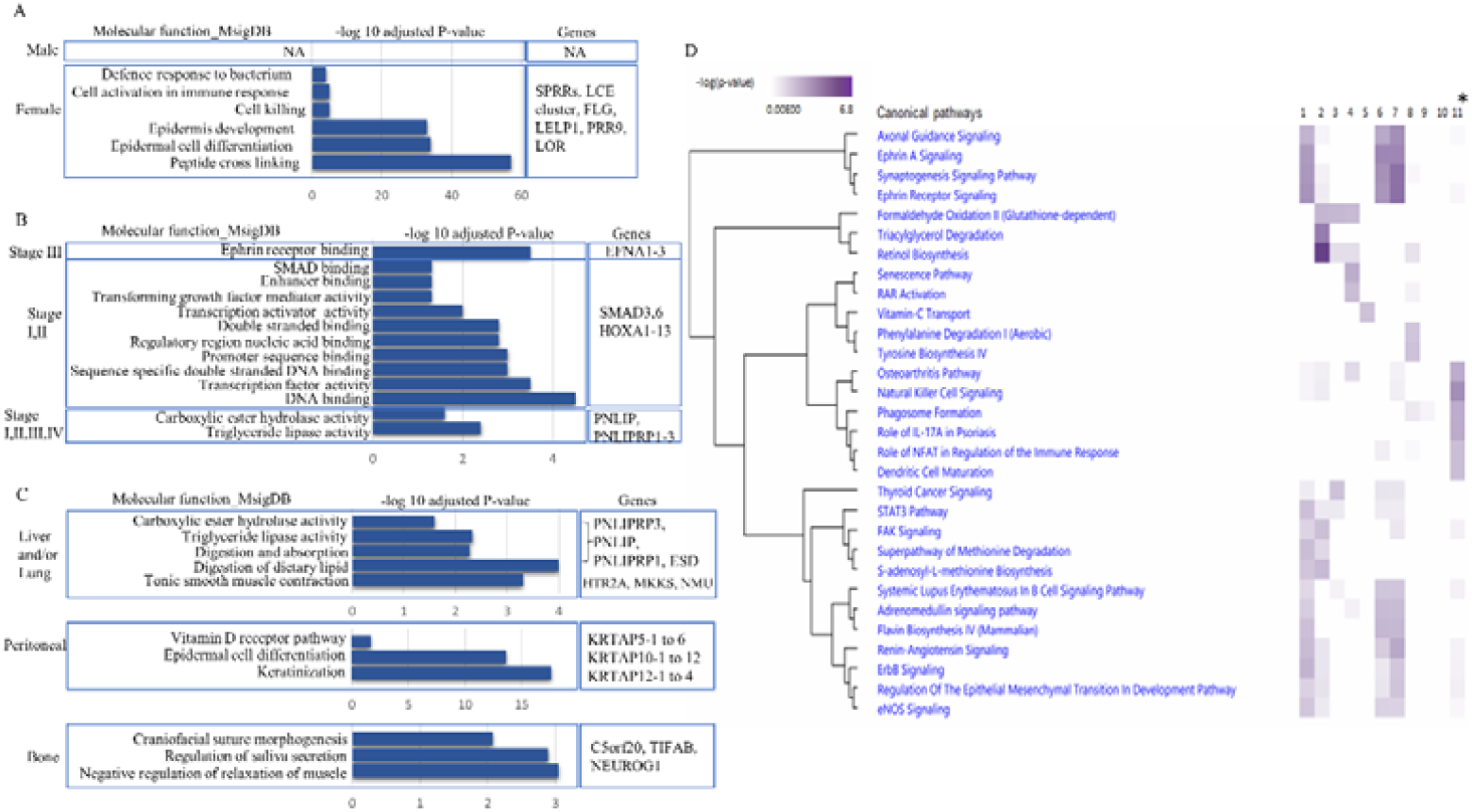
Distinct molecular functions and pathways associated with metastasis in different disease subtypes. Molecular functions from MsigDB investigation of metastasis-positive vs metastasis-negative cases stratified by A) gender; B) left-sided tumor stages and C) left-sided distal organ-specific metastasis. D) Canonical pathways from IPA interrogation of metastasis-positive vs metastasis-negative cases in various disease subtypes. Column 1= all metastasis cases; 2 = left-sided (left colon and rectum) metastasis; 3 = Stage I/II metastasis; 4 = left-sided Stage I/II metastasis; 5 = Stage III metastasis; 6 = left-sided Stage III metastasis; 7 = left-sided liver and/or lung metastasis in Stage I/II/III; 8 = bone-metastasis; 9 = left-sided peritoneal metastasis; 10 = male metastasis; *11 = female metastasis. Metastasis cases were compared to corresponding metastasis-negative cases of the overall or disease subtype. Only significant pathways with -log10p>2 was represented.

### 3.5. Pathway analysis with IPA

The candidate genes identified with FUMA were subsequently imported into the IPA software for further analysis. Since the right-sided colon cancer did not yield significant genesets, this subtype was not profiled.

Thirty different canonical pathways clustered according to their similarity were identified (Figure 3D). These pathways ranged from axonal guidance signaling to eNOS signaling. The disease subtypes with significant pathways closest to all cases were the left-sided liver and/or lung metastasis in stages I/II/III, followed closely by left-sided stage III metastasis. When right-sided colon metastasis cases were omitted, the left sided metastasis have higher significance in the cluster comprising the glutathione-dependent formaldehyde oxidation, triacylglycerol degradation and retinol bio-synthesis pathways. The glutathione-dependent formaldehyde oxidation pathway was also significant in stages I/II metastasis but absent in stage III metastasis cases. Left-sided stage I/II metastasis also has significant senescence and RAR activation pathways. Bone metastasis cases have significant canonical pathways in the cluster comprising phenylalanine degradation and tyrosine biosynthesis which were not found in other disease subtypes. Notably, female metastasis cases (lane 11 – Figure 3D) have a significant unique cluster comprising six pathways ranging from osteoarthritis pathway, natural killer cell signaling, phagosome formation, IL-17 activation in psoriasis, NFAT regulation to dendritic cell maturation that were not significant in the male metastasis cases.

### 3.6. Replication of the top SNPs from the GWAS panel in RP

We selected 23 SNPs with association testing -log10p-values ≥ 5.0 from the overall as well as subtype datasets to replicate in the RP (Supplemental Table S3). These SNPs were genotyped using the Fluidgm SNPtype arrays. Two SNPs rs2131902 and rs7711080 at chromosomes 1q21.3 and 5q34 respectively had increased significance in the pooled GWAS and RP analysis although they have not reached genome-wide significance of 7.3 probably due to the small RP size (Table 2). Notably, for rs2131902, pooled analysis for the lung and or liver metastasis subtype has higher significance than that of the overall (stage I-IV) analysis indicating that other metastasis-positive cases (such as peritoneal or bone metastasis) were probably confounders. One other SNPs rs2919408 at chromosomes 8q13.3 was validated for left-sided bone-metastasis (Table 2). Sample sizes of various disease subtypes are represented in Supplemental Table S4.

## 4. Discussion

Accumulating evidence indicate that the germline plays an important role in shaping the tumor genome [11-14]. Indeed, common germline variants could impact somatic alterations and lead to earlier tumor development and progression [14]. In this study, we adopted the case-case GWAS approach to interrogate whether there are germline metastasis risk variants contributing to distal-organ metastasis, the major cause of CRC mortality. Our inclusion criteria of the cases were stringent to ensure validity and reproducibility of the results.

We started with about 3,000 well-defined cases that were eventually dwindled down to 2677 by both clinical and genotyping quality filter exclusion. In addition, we have another 837 cases filtered by the same criteria as the independent RP. There was no significant difference in the clinicopathological features of the metastasis-positive vs metastasis-negative cases except for tumor stage, recapitulating that early-stage CRCs and stage III CRC have diverse metastasis rate (Table 1). Importantly, metastasis-positive and metastasis-negative cases were represented almost equally in both the GWAS panel (1282 metastasis-positive vs 1395 metastasis-negative) and the RP (346 metastasis-positive vs 491 metastasis-negative).

We identified several regions of interest (ROI) comprising SNPs with -log10 p ≥ 5 in various chromosomes through genome-wide association testing using the additive model in both the overall dataset as well as the various disease subtypes (Figure 1; Supplemental Table S1; and Supplemental Table S2). Of note, these ROIs were different from those identified by case-control GWAS for disease occurrence suggesting that the risk variants affecting disease etiology were different from those for metastasis (https://www.ebi.ac.uk/gwas/). It further suggests that “collider bias” will not be an issue should these metastasis risk variants be used in any mendelian randomization studies in future [15, 16]. These data were substantiated by a recent study which reported that none of the CRC risk variants from case-control GWAS were associated with survival [17]. In contrast, we found expression of two of the five metastasis-relevant genes profiled significantly associated with metastasis-free survival (see discussion below). Further, these metastasis risk SNPs have larger effective sizes (OR>1.2) than the disease occurrence risk variants (OR≤1.2). Thus, genetic predisposition may play a greater role in metastasis than in cancer etiology [18].

Candidate genes within 500 kb of these SNPs were previously implicated in different metastasis steps, ranging from EMT, mechanical stress in systemic circulation to implantation at distal organ site. This reaffirmed that metastasis is a multi-step process [19, 20]. For rs16889360, it is so specific for bone-metastasis that in the overall analysis, the -log10 p-value of the association test is only 2.06 while in the bone-metastasis vs metastasis-negative analysis, it has risen to 7.97, despite a drastic 88% drop in metastasis-positive cases (Figure 1B; Supplemental Table S2). Such specific risk variants also have large effect sizes (OR=3.6) suggesting that they are probably rare variants that have evolved recently and hence likely to be population specific [21].

We chose 23 top ranking SNPs from association testing of the GWAS panel to replicate in the RP. Only 3 of the SNPs have increased significance in the pooled GWAS and RP analysis. rs7711080 appeared to be tagging all cases (Stage I-IV) while rs2131902 and rs2919408 tagged causal variants in lung/liver and bone metastasis respectively (Table 2). rs2131902 at chromosome 1q21.3 is a protective allele (odds ratio <1 compared to major allele) while the other two are risk alleles. While the rest of the SNPs could be false positives (‘winners curse’) (Supplemental Table S3), accumulating evidence has indicated that single SNP analysis may not be optimal to identify disease-relevant genes for more complex genetic models such as epistasis and pleiotropy [22, 23]. Moreover, trait-associated loci are often driven by multiple genetic variants acting together [24]. Recent studies leveraged on computational methods to prioritize candidate target genes from risk variants using multiple resources such as chromatin immunoprecipitation, sequencing and transcriptomics data [25, 26].

We imported risk SNPs with -log10 p ≥ 5 into FUMA for further functional annotation (Supplemental Figure S2). FUMA is a web-based application that has been widely used recently and yielded many biologically relevant genesets for various diseases [27, 28]. Interestingly, analysis of the overall dataset revealed that the top hallmark geneset is glycolysis with 5 genes (HAX1, HMMR, EFNA3, SRD5A3 and TGFA) from 4 genomic risk loci implicated. Although metabolic reprogramming is known to be involved in sporadic CRC etiology, less is known of its role in metastasis [29, 30]. We confirmed by qRT-PCR analysis that the expression of two of the genes, HMMR and HAX1, were significantly associated with metastasis-free survival and that the two genes appeared to be acting synergistically (Figure 2). HMMR is a transmembrane receptor that also functions internally, as a microtubule-associated protein that plays a critical role in progression and proliferation of various cancers [31]. Some studies have indicated that overexpression of HMMR was significantly correlated with worse survival in various cancers [32-34]. However, a recent review has shown that both over and under-expression of HMMR maybe associated with aggressive tumors thus its role in tumorigenesis could be context-dependent [31]. HAX1 is an adaptor protein that regulates cell migration, invasion, and angiogenesis [35].

HAX1 is localized to the inner mitochondria membrane and plays an important role in bioenergetics [36]. In HAX1-deficient mitochondria, protein synthesis is elevated and hence, TCA and glycolysis cycles are significantly impacted. HMMR and HAX1 are located about 300 kb from the two SNPs with increased significance in the pooled GWAS and RP analysis, rs7711080 and rs2131902 respectively, suggesting that these two genes are the functional genes tagged by the two SNPs. To our knowledge, HAX1 has not been previously reported to act synergistically with HMMR in impacting metastasis-free survival in CRC. A recent study has reported that energy production is higher in metastases compared to primary tumors [37]. Our data suggest that metabolic reprogramming in primary tumors prone to metastasis could have proceeded via perturbation of these genes. The expression of the other 3 transcripts were not significantly different between metastasis-positive and metastasis-negative cases tested. Their expression could be perturbed at the proteomics or metabolomics level.

Of note, the significant molecular functions and pathways for female metastasis-positive cases were very different from the male metastasis-positive cases despite sex chromosomes being excluded from the genome-wide analysis (Figure 3A and 3D). The six canonical pathways in the same cluster (lane 11, Figure 3D) such as the natural killer cell signaling, were immune-related pathways, substantiating previous evidence that there is significant gender-differences in immune response and cancer etiology [38, 39]. Our data indicate further that immune-mediated mechanisms may also contribute significantly to metastasis risk for female CRC patients.

The molecular functions and pathways associated with metastasis from left-sided early stage I/II CRC were also very different from those of left-sided stage III CRC (Figure 3B and lanes 4 and 6 Figure 3D). The genes (HOXA1-13, SMAD3,6) and crosstalk between the senescence and retinol acid receptor (RAR) activation pathways indicated that the EMT process, which initiate metastasis, was more important in early stage-than in stage III-CRC, reaffirming that different pathways are involved in different metastasis stages [40, 41]. HOX proteins are master transcription factors occupying the top of many regulatory networks that induce EMT in various cancers [42, 43]. Ephrin receptor binding or signaling was predominantly active in left-sided stage III metastasis-positive cases (Figure 3B and lane 6, Figure 3D). It is also a significant canonical pathway in the overall and left-sided liver and/lung (Stage I-III) datasets (lanes 1 and 7, Fig. 3D) respectively. Ephrin receptors were the most abundant receptor tyrosine kinase in mammals and upon binding of the ligands, effected downstream signaling such as RAS to increase cell proliferation. Pathways in this cluster also influence axonic guidance, cell migration, angiogenesis and synaptic plasticity [44]. This suggests that ephrin receptor signaling and associated pathways were the significant pathways associated with metastasis risk overall, although these pathways were not as significant in subtypes such as stage I/II metastasis (Fig. 3B and 3D). It is intriguing that CRC metastasis risk was closely correlated with these pathways which also play important role in neurotransmission and nervous system signaling thus implying that CRC metastasis and neuro-dysfunction may share common predisposition. Indeed, recent epidemiological evidence has alluded to an inverse co-morbidity relationship between cancer and neurodegenerative diseases [19, 45-46]. The role of the Eph/ephrins in cancer metastasis and as potential therapeutic targets were summarized in recent reviews [47, 48].

The most significant pathways associated with left-sided metastasis from all tumor stages (Stage I-IV) were the retinol biosynthesis and triacylglycerol degradation pathways suggesting that free radical scavenging and lipid metabolism were important contributors to left-sided colon and rectal metastasis (Figure 3B and lane 2 Figure 3D) [49]. Triglyceride catabolism by carboxylesterase promoted aggressiveness in CRC [50]. A recent epidemiological study has also identified the carboxylic ester and the triglyceride metabolites as important risk factors in East Asian CRC patients [51], thus sup-porting the findings of this study that perturbation in lipid metabolism contributed significantly to CRC metastasis. This perturbation is likely mediated by lipases such as the PNLIPRPs in lipid droplets which accumulated in cancer cells undergoing metabolic stress [52, 53].

Interrogating distal organ-specific datasets revealed that left-sided liver and/or lung metastasis-positive cases have similar molecular functions (triglyceride lipase and carboxylic ester hydrolase activities) as that of left-sided colon and rectal metastasis. This was internally consistent as this subtype contributed the most (92.7%) metastasis cases to the metastasis from left-sided CRC (Figure 3C). The molecular functions high-lighted in left-sided peritoneal and bone metastasis respectively were however very different from that of the liver and lung metastasis-positive cases (Figure 3C) suggesting that some distal-organ specific tumor microenvironmental factors were probably important contributors to the metastasis to these organs (Figure 3C). While there is literature linking vitamin D pathways to metastasis of various cancers [54, 55], the association of the keratinization geneset to left-sided CRC peritoneal metastasis appeared to be novel. To our knowledge, the CRC-associated peritoneal metastasis in our panel is the largest series (n =206) for East Asian thus far. Similarly, there are reports linking the implicated bone metastasis genes to craniofacial suture morphogenesis and muscle regulation respectively [56, 57]; the relationship with regulation of saliva secretion however is less clear (Figure 3C).

Previous studies investigating genetic risk in relation to metastatic cancers were based mostly on the candidate gene approach and tumor staging was often used as surrogate endpoint for metastasis [58, 59]. This is not ideal as early-stage (stage I/II) CRCs can still succumb to metastasis and as shown in this study, the metastasis initiation pathways in early-stage CRCs were distinct from pathways associated with lymph-node involved stage III CRCs. Further, many studies made use of TCGA and GEO public databases which lack detailed clinical information [60]. In recent years, case-control GWAS studies have uncovered many common genetic risks variants for cancer initiation (https://www.ebi.ac.uk/gwas/) but not for metastasis per se due likely to the difficulty of defining metastasis accurately. The strength of this study lies in the careful selection of almost equal number of metastasis-positive and metastasis-negative cases by clinical definition. This study is probably the first to use clinical criteria of metastasis to uncover metastasis-relevant genes for various CRC subtypes through case-case GWAS, subsequent in silico analysis followed by validation at the tissue level. The limitation is the relatively small sample size for the initial case-case GWAS study.

## 5. Conclusions

The results of this study reaffirm that CRC is a very heterogeneous disease and that disease-subtype specific metastasis risk genes would need to be included as features in the development of subtype-specific (for instance tumor stage- or gender-specific) metastasis-risk signatures for clinical application. We have previously developed metastasis-prone signature with high positive predictive values by machine learning implementation [8]. The validation of subtype-specific metastasis-risk genes as shown by the workflow of this study would enable the development of subtype specific metastasis signatures which could aid in early identification of stage I-III CRC patients who are prone to metastasis for tailored management instead of a one-size-fits-all approach [61].

## Data Availability

Supplementary data are attached in additional files. The other datasets used and/or analyzed during the current study are available from the corresponding author on reasonable request.

## Supplementary Materials

File S1. Supplementary Methods for genome-wide genotyping, genotyping calling and quality control steps, FUMA SNP2GENE process and qRT-PCR assay conditions; Figure S1. PCA and Q-Q plots; Figure S2. Combined workflow chart; Figure S3. FUMA output for overall dataset analysis; Table S1. Region of Interest from association testing of overall dataset; Table S2. Candidate loci from analysis of subtype datasets; Table S3. 23 SNPs replicated in Replication Panel; Table S4. Sample sizes of overall and disease subtype datasets.

## Author Contributions

Conceptualization, Iain Tan and Peh Yean Cheah; Data curation, Lai Fun Thean, Michelle Wong and Michelle Lo; Formal analysis, Lai Fun Thean, Fei Gao and Peh Yean Cheah; Funding acquisition, Peh Yean Cheah; Investigation, Lai Fun Thean, Michelle Wong and Michelle Lo; Methodology, Lai Fun Thean and Peh Yean Cheah; Project administration, Lai Fun Thean, Michelle Lo and Peh Yean Cheah; Resources, Iain Tan, Evelyn Wong, Emile Tan and Choong Leong Tang; Supervision, Peh Yean Cheah; Validation, Lai Fun Thean, Michelle Wong and Michelle Lo; Writing – original draft, Peh Yean Cheah; Writing – review & editing, Lai Fun Thean and Peh Yean Cheah.

## Institutional Review Board Statement

The study was conducted in accordance with the Declaration of Helsinki and approved by SingHealth Central Institutional Review Board (CIRB # 2018/2631).

## Informed Consent Statement

Informed consent was obtained from all subjects involved in the study.

## Data Availability Statement

Supplementary data are attached in additional files

The other datasets used and/or analyzed during the current study are available from the corresponding author on reasonable request.

## Acknowledgments

We thank all patients who provided materials for the study.

