## Supplemental File S1 for "Functional annotation with expression validation identifies novel metastasis-relevant genes from post-GWAS risk loci in sporadic colorectal carcinomas"

**Supplementary Methods**

Genome-wide genotyping

Genomic DNA was extracted using the Qiagen DNeasy blood and tissue kit according to the manufacturer’s protocol. Whole genome scan was performed with Affymetrix^TM^ Genome-Wide Human SNP Array 6.0 featuring 906,000 SNPs and 946,000 copy number probes. A total of 600ng of DNA was digested with the restriction enzymes, NspI and StyI, followed by adaptor ligation and amplification, fragmentation, biotin end-labeling, hybridization, washing and staining on a 4-module Affymetrix Fluidic Station. The samples were randomized, and each run has both metastatic-positive and metastatic-negative samples. Scanning was performed with the Affymetrix 3000-7G scanner.

Genotyping calling and quality control steps

Genotype calls and quality controls including the Contract Quality Control (CQC) metric were performed using the Genotyping Console 4.0 software. CQC predicts the genotyping performance and measures the separation of allele intensities by transforming A and B values into a contrast value with a given subset of SNPs, with A and B being the median intensities of replicate probes for the A and B alleles of a SNP. CQC analysis was performed for each batch of samples and mean of the CQC was calculated for samples that passed the 0.4 threshold. Samples with a CQC <0.4 were either repeated or excluded when the repeated sample still did not pass the CQC requirement. The mean passing CQC is >1.7.

The Analysis Power Tools (APT) analyse Affymetrix microarray data focusing on CEL files. Experimental qualities for the SNP6 array were assessed with apt-geno-qc program that uses the model-based algorithm DM. Report file containing the QC values for each CEL files was created. The apt-probeset-genotype program that implements BRLMM-P was applied to make genotype calls with 1-dimensional clustering. The per-sample QC metric of at least 86% QC call rate was set as the inclusion criteria when performing genotype calling.

SNP quality check was performed with three components of SNPolisher under APT: ps-metrics, ps-classification and OTV (off target variants) caller. Ps-metrics generated SNP QC metrics to identify problematic SNPs. The ps-classification program classified SNPs into seven categories based on the QC metrics generated by ps-metrics: PolyHighResolution, MonoHighResolution, OTV, CallRateBelowThreshold, NoMinorHom, Hemizygous and Other. Based on the classification, the output file contained the recommended SNP list for further downstream analysis. OTV refers to SNPs with the genotype sequences showing inconsistency compared to the hybridization probes, and with low hybridization intensities. OTV caller performed post-processing analysis on the potential OTVs identified under ps-classification and created another list of true OTVs. The apt-package-util and apt-probeset-genotype were performed to create CHP files of samples that qualified for further analysis.

FUMA SNP2GENE process

GWAS summary data for about 647,545 SNPs were uploaded onto FUMA SNP2GENE for post-GWAS analyses. To define genomic loci of metastasis risk, pre-calculated linkage disequilibrium (LD) structure based on 1000G phase 3 East Asian population was used. Independent significant SNPs with genome wide *P* ≤ 1E-05 and pairwise r^2^ < 0.7 were identified. All SNPs with r^2^ ≥ 0.7 with the identified independent significant SNPs were included for subsequent annotations and gene prioritization. Lead SNPs were defined by clumping of independent significant SNPs with pairwise SNPs r^2^ < 0.1. The threshold value of *P* <0.05 was applied to candidate SNPs in linkage disequilibrium (LD) with independent significant SNPs. Distance of LD blocks of independent significant SNPs to be merged into a single genomic locus was 250kb. Positional mapping was performed based on ANNOVAR annotations with the physical distance of 10kb.

qRT-PCR assay conditions

The PrimePCR™ Probe Assays for the gene of interests (GOI) labelled with FAM were HMMR (assay ID: qHsaCEP0039054), SRD5A3 (assay ID: qHsaCEP0051465), TGFA (assay ID: qHsaCEP0053322), EFNA3 (assay ID: qHsaCEP0052409) and HAX1 (assay ID: qHsaCEP0040572). The PrimePCR™ Control Assay was ACTB labelled with HEX (assay ID: qHsaCEP0036280). The 10µl duplex qPCR reaction mix contained 5µl of 20X SsoAdvanced™ Universal Probes Supermix (Bio-Rad), 0.5µl of probe assay and 0.5µl of control assay, 1µl cDNA (25ng) and 3µl of nuclease-free water. The PCR condition was initial denaturation at 95°C 4min, followed by 40 cycles of denaturation at 95°C 3s and annealing and extension at 62°C 30s.
