## Supplemental Figure S1 for "Functional annotation with expression validation identifies novel metastasis-relevant genes from post-GWAS risk loci in sporadic colorectal carcinomas"

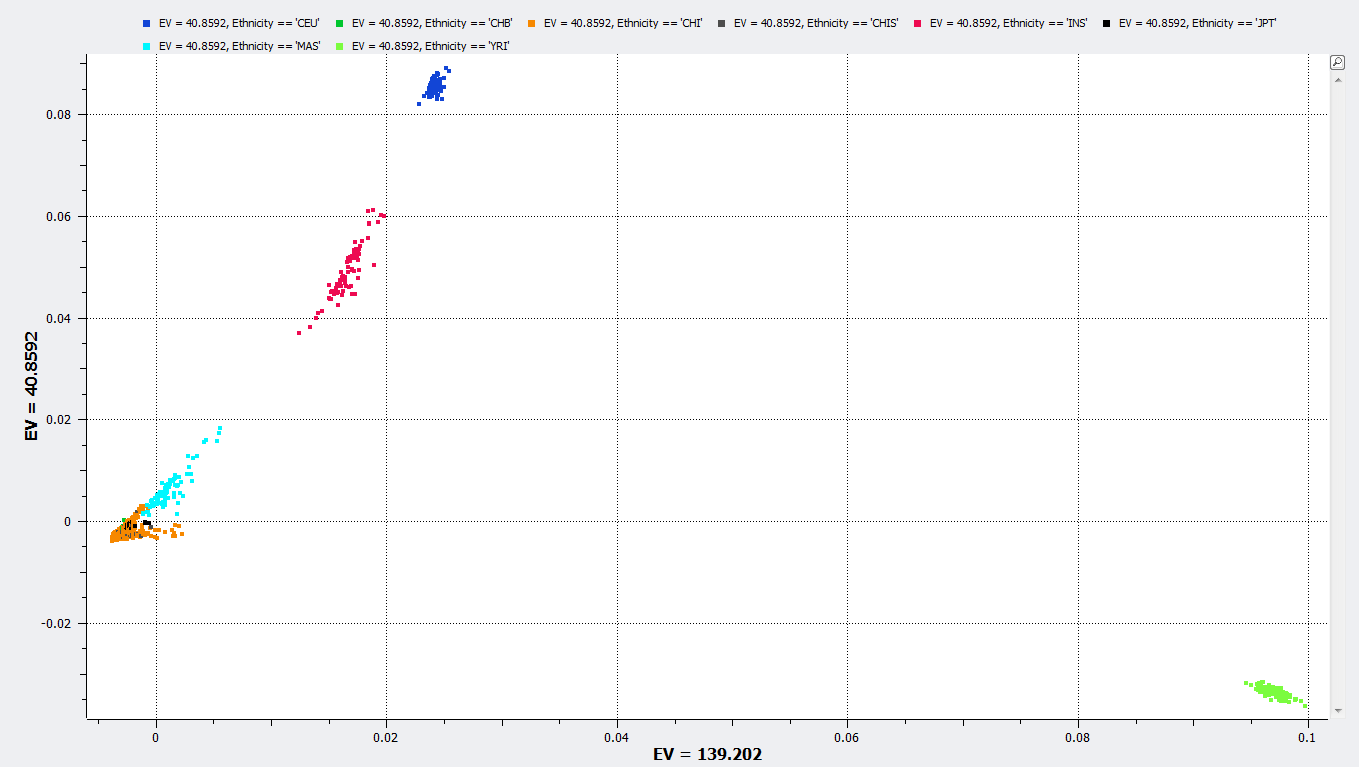

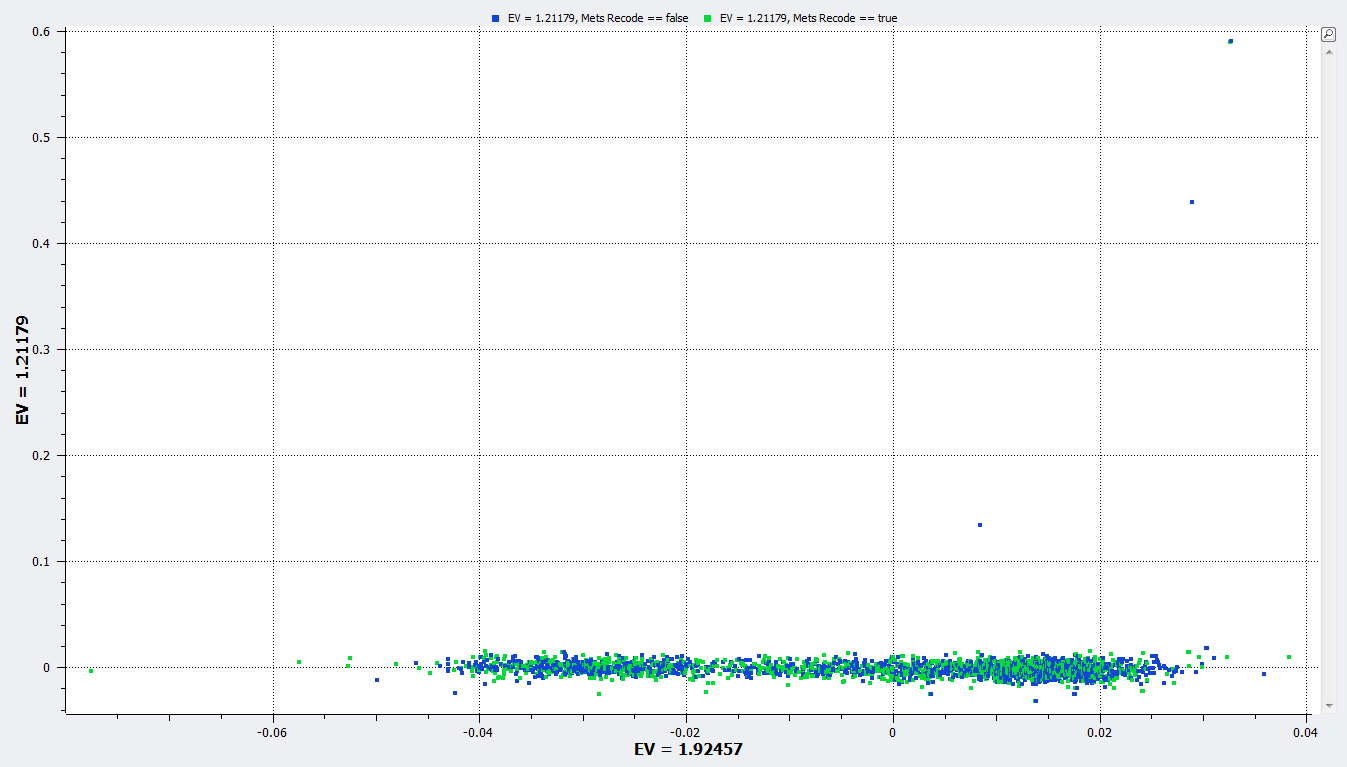

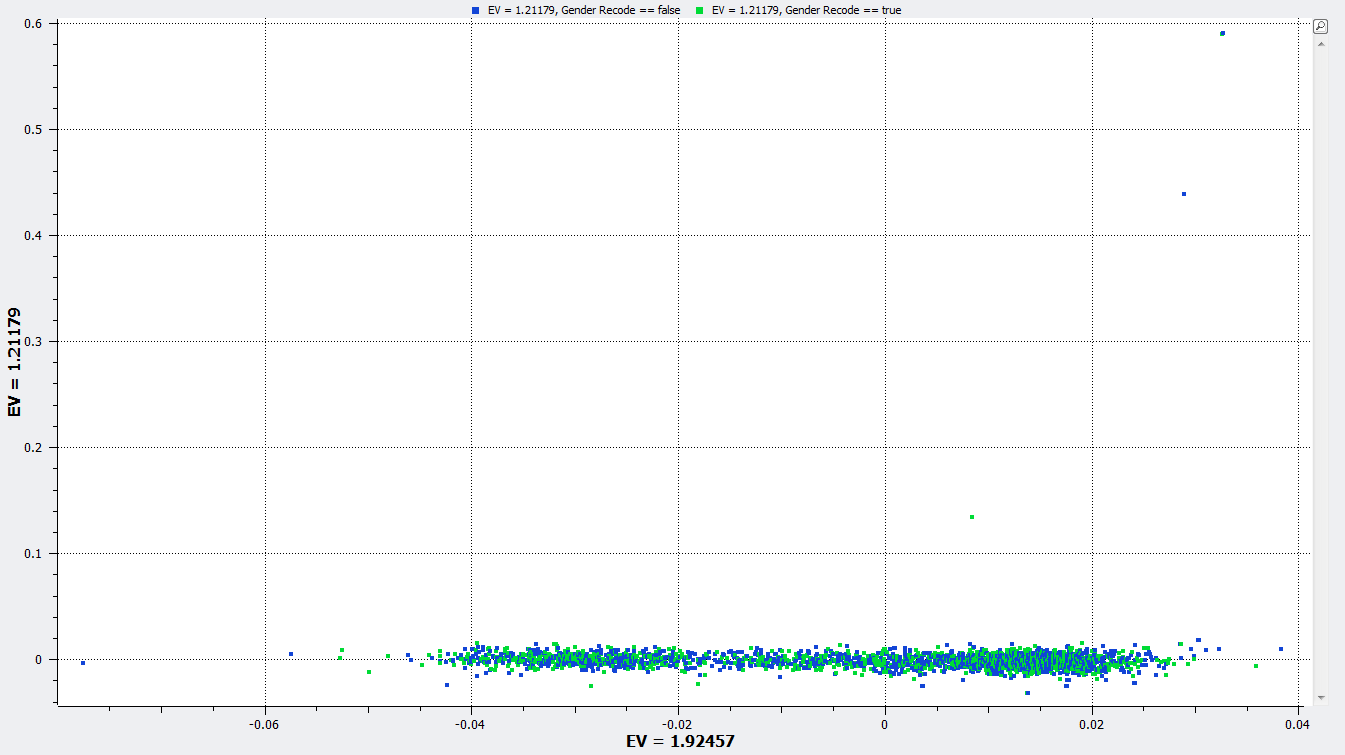

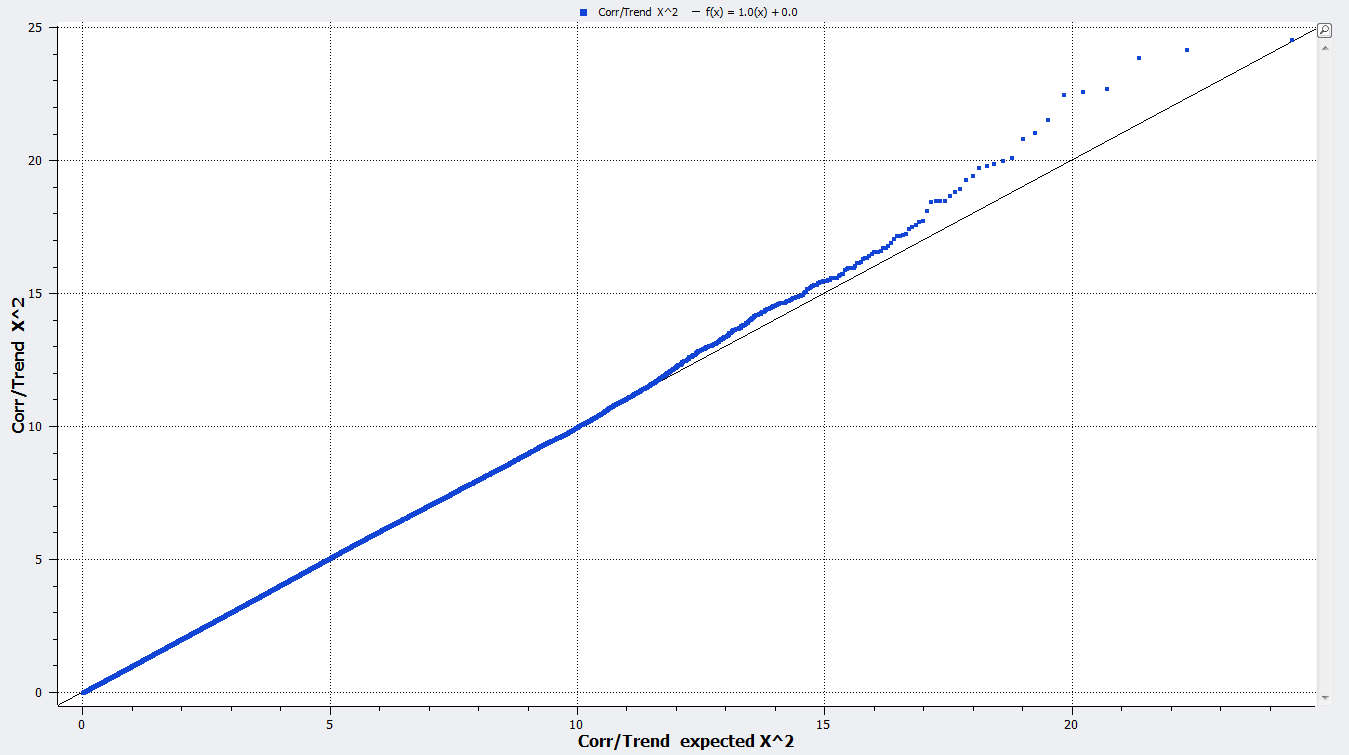

A

B

C

D

**Fig. S1**: A) PCA plot of 4191 samples including our genotyped samples, the SGVP and the HapMap subjects. Arrow indicates all our samples clustered with the Chinese subjects (orange) in SGVP and HapMap; B) PC1 (x-axis) vs PC2 plot (y-axis) for metastasis-positive (green) and metastasis-negative (blue) cases; C) PC1 vs PC2 plot for male (green) vs female (blue)cases; D) Quantile-Quantile plot (λ=1.003).
