## Supplemental Figure S2 for "Functional annotation with expression validation identifies novel metastasis-relevant genes from post-GWAS risk loci in sporadic colorectal carcinomas"

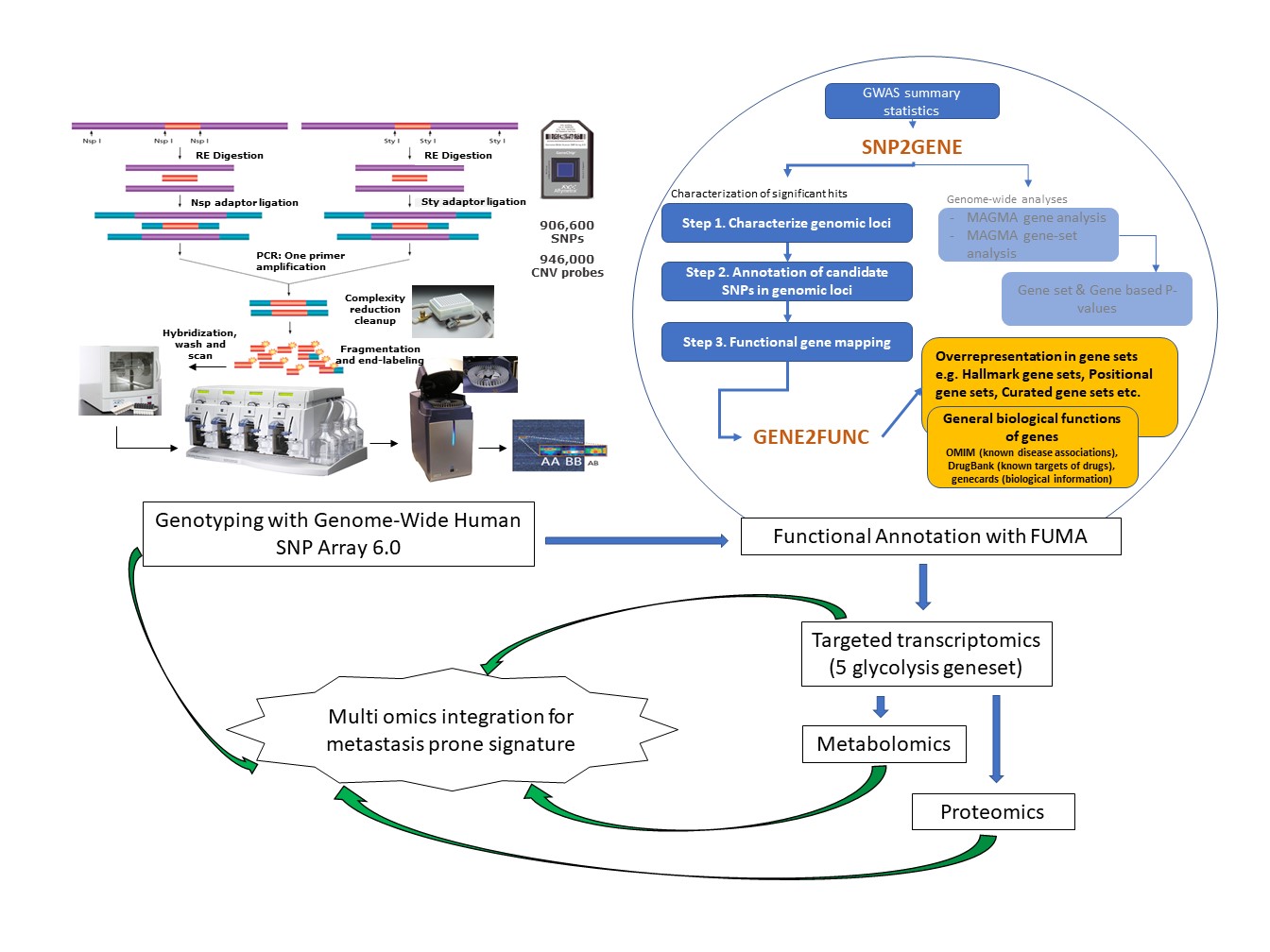

**Fig.S2**: Combined genome-wide SNP analysis and functional annotation with expression validation identified metastasis-relevant genes
