## Supplemental Figure S3 for "Functional annotation with expression validation identifies novel metastasis-relevant genes from post-GWAS risk loci in sporadic colorectal carcinomas"

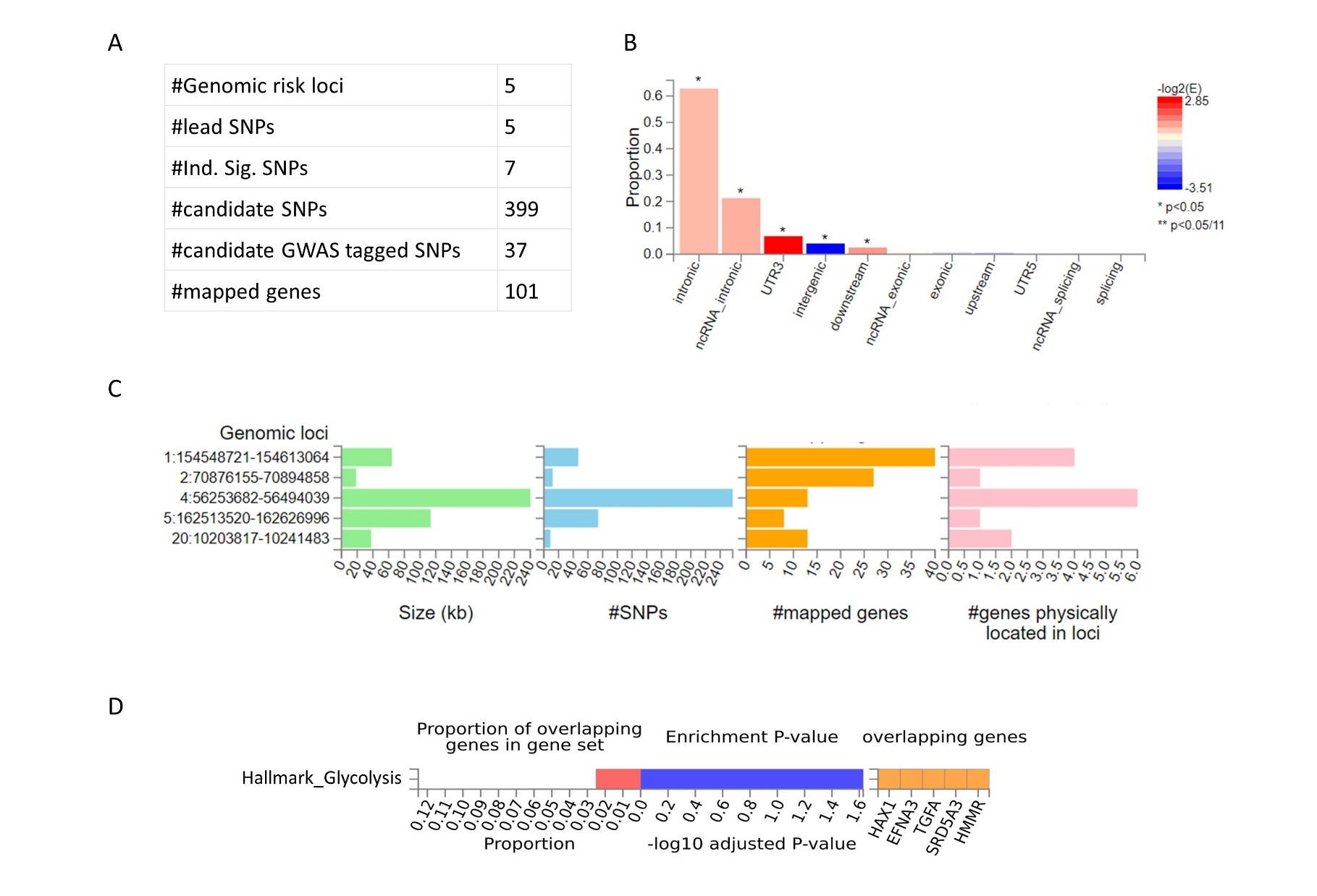

**Fig.S3**: FUMA output for overall dataset analysis. A) Summary of SNPs and mapped genes; B) Functional consequences of SNPs on genes; C) Summary per genomic risk locus and D) Glycolysis geneset and genes identified in Gene2Func workflow.
