## Supplemental Table S1 for "Functional annotation with expression validation identifies novel metastasis-relevant genes from post-GWAS risk loci in sporadic colorectal carcinomas"

**Table S1: Region of Interest from association testing of overall dataset**

| **Chromosome** | **Corr/Trend -log10 P (top SNP)** | **Total # of SNPs -log10 P >5.0 (#SNP 4.0-4.4)** | **OR (95% CI)** |
| --- | --- | --- | --- |
|  |  |  | **Dd vs dd** |
| 20p12 | 6.13 | 9 (2 SNPs 4.0-4.4) | 1.36 (1.16, 1.60) |
| 5q34 | 5.97 | 8 | 1.48 (1.23, 1.78) |
| 2p13.3 | 5.35 | 2 | 1.45 (1.19, 1.72) |
| 4q12 | 5.11 | 1 (2 SNPs ~4.1) | 0.74 (0.63, 0.88) |
| 1q21 | 5.08 | 2 (3 SNPs 4.0-4.4) | 0.81 (0.69, 0.96) |

Overall [stages I/II/III (metastasis-positive) and IV cases vs stages I/II/III (metastasis-negative cases)]; Dd heterozygous minor-major allele; dd homozygous major allele
