## Supplemental Table S2 for "Functional annotation with expression validation identifies novel metastasis-relevant genes from post-GWAS risk loci in sporadic colorectal carcinomas"

**Table S2:** **Candidate loci from analysis of subtype datasets**

| **SNP ID** | **Corr/Trend -log10 P** | **Corr/Trend P** | **Odds Ratio (95% CI)** | **Subtype Analysis** | **Total, *N_T_*** | **Mets -ve, *N_0_*** | **Mets +ve, *N_1_*** | **MAF*** | |
| --- | --- | --- | --- | --- | --- | --- | --- | --- | --- |
|  |  |  |  |  |  |  |  | **(Mets -ve)** | **(Mets +ve)** |
| rs363020 | 6.13 | 7.36E-07 | 1.36 (1.16, 1.60) | Overall | 2677 | 1395 | 1282 | 0.19 | 0.25 |
|  | 6.86 | 1.37E-07 | 1.59 (1.29, 1.96) | Rectum + LT Colon stage I/II/III | 1778 | 1141 | 637 | 0.19 | 0.26 |
| rs7711080 | 5.97 | 1.06E-05 | 1.48 (1.23, 1.78) | Overall | 2677 | 1395 | 1282 | 0.45 | 0.52 |
|  | 7.05 | 8.91E-08 | 1.68 (1.36, 2.06) | Rectum + LT Colon | 2147 | 1120 | 1027 | 0.44 | 0.53 |
| rs233268 | 1.53 | 2.93E-02 |  | Overall | 2677 | 1395 | 1282 | 0.21 | 0.24 |
|  | 6.93 | 1.19E-07 | 1.76 (1.28, 2.41) | Peritoneal mets vs mets -ve (Rectum + LT Colon) | 1326 | 1120 | 206 | 0.21 | 0.33 |
| rs16889360 | 2.06 | 8.66E-03 |  | Overall | 2677 | 1395 | 1282 | 0.027 | 0.04 |
|  | 7.97 | 1.07E-08 | 3.66 (2.18, 6.14) | Bone mets vs mets -ve | 1545 | 1395 | 150 | 0.028 | 0.094 |
| rs1491041 | 0.14 | 7.22E-01 |  | Overall | 2677 | 1395 | 1282 | 0.097 | 0.096 |
|  | 6.59 | 2.60E-07 | 2.00 (1.33, 3.02) | Bone mets vs mets -ve (Rectum + LT Colon) | 1257 | 1124 | 133 | 0.09 | 0.2 |

***MAF** Minor allelic frequency
