## Supplemental Table S3 for "Functional annotation with expression validation identifies novel metastasis-relevant genes from post-GWAS risk loci in sporadic colorectal carcinomas"

**Table S3:** **23 SNPs replicated in Replication Panel**

| **rsID** | **Analyses** | **GWAS** | | | | | **GWAS + RP** | | | | | | |
| --- | --- | --- | --- | --- | --- | --- | --- | --- | --- | --- | --- | --- | --- |
|  |  | **N** | **Cases** | **Controls** | **Corr/Trend -log10 P** | **Corr/Trend P** | **N** | **Cases** | **Controls** | **Corr/Trend -log10 P** | **Corr/Trend P** | | **OR** |
| rs363020 | I/II/III/IV | 2677 | 1282 | 1395 | 6.13 | 7.36E-07 | 3514 | 1628 | 1886 | 5.44 | 3.62E-06 | | 1.31 (1.14, 1.51) |
| rs7711080 | Rectum + LT Colon | 2147 | 1027 | 1120 | 7.05 | 8.91E-08 | 2796 | 1295 | 1501 | 6.79 | 1.62E-07 | | 1.59 (1.33, 1.91) |
|  | I/II/III/IV | 2677 | 1282 | 1395 | 5.97 | 1.06E-06 | 3514 | 1628 | 1886 | 6.79 | 1.62E-07 | | 1.45 (1.24, 1.71) |
| rs2131902 | Liver +/or Lung mets vs no mets stage II/III | 1806 | 632 | 1176 | 6.49 | 1.93E-07 | 2351 | 778 | 1573 | 7.51 | 3.08E-08 | | 0.76 (0.64, 0.91) |
|  | Liver +/or Lung mets vs no mets stage I/II/III | 2037 | 644 | 1389 | 6.70 | 5.76E-07 | 2676 | 799 | 1877 | 6.60 | 2.52E-07 | | 0.80 (0.67, 0.95) |
|  | Liver +/or Lung mets vs no mets | 2480 | 1085 | 1395 | 6.10 | 7.92E-07 | 3224 | 1359 | 1885 | 6.86 | 1.38E-07 | | 0.82 (0.71, 0.95) |
|  | II/III | 1942 | 766 | 1176 | 6.08 | 8.24E-07 | 2535 | 962 | 1573 | 7.28 | 5.29E-08 | | 0.75 (0.63, 0.89) |
| rs7326071 | Peritoneal mets vs mets -ve (Rectum) | 712 | 92 | 620 | 6.72 | 1.91E-07 | 933 | 108 | 825 | 4.36 | 4.34E-05 | | 4.10 (2.08, 8.06) |
| rs12500837 | Rectum + LT Colon | 2147 | 1027 | 1120 | 5.81 | 1.53E-06 | 3514 | 1628 | 1886 | 3.81 | 0.000156274 | | 0.73 (0.62, 0.87) |
| rs10756451 | Liver mets vs non-liver mets (Rectum + LT Colon) | 1032 | 636 | 396 | 6.39 | 4.07E-07 | 1337 | 801 | 536 | 3.39 | 0.00040436 | | 0.64 (0.50, 0.83) |
| rs17689224 | Liver mets vs non-liver mets (LT Colon + RT Colon) | 674 | 440 | 234 | 6.33 | 4.72E-07 | 875 | 548 | 327 | 6.63 | 2.33E-07 | 0.28 (0.17, 0.48) | |
| rs10953868 | LT Colon stage ABD | 580 | 271 | 309 | 6.07 | 8.49E-07 | 882 | 394 | 488 | 3.52 | 0.000301694 | | 1.59 (1.146, 2.17) |
| rs725050 | Rectum + LT Colon stage I/II/IV | 1272 | 593 | 679 | 6.87 | 1.35E-07 | 1645 | 756 | 889 | 5.78 | 1.65E-06 | | 1.56 (1.27, 1.93) |
| rs7943117 | LT Colon stage I/II/IV | 580 | 271 | 309 | 6.29 | 5.18E-07 | 735 | 337 | 398 | 6.59 | 2.57E-07 | | 0.48 (0.35, 0.66) |
| rs233268 | Peritoneal mets vs mets -ve (Rectum + LT Colon) | 1326 | 206 | 1120 | 6.93 | 1.19E-07 | 1759 | 258 | 1501 | 4.56 | 2.78E-05 | | 1.39 (1.05, 1.84) |
| rs1261225 | Peritoneal mets vs mets -ve (LT Colon) | 614 | 114 | 500 | 7.08 | 8.24E-08 | 831 | 151 | 680 | 6.44 | 3.66E-07 | | 4.78 (2.48, 9.20) |
| rs10844418 | Peritoneal mets vs mets -ve (Rectum) | 712 | 92 | 620 | 6.24 | 5.74E-07 | 933 | 108 | 825 | 5.51 | 3.09E-06 | | 4.00 (2.08, 7.67) |
| rs12973240 | Lung mets vs non-lung mets (overall) | 1282 | 620 | 662 | 6.52 | 3.05E-07 | 1628 | 742 | 886 | 6.06 | 8.77E-07 | | 0.50 (0.37, 0.68) |
| rs969095 | Bone mets vs mets -ve (Overall) | 1545 | 150 | 1395 | 7.71 | 1.95E-08 | 2057 | 171 | 1886 | 6.55 | 2.83E-07 | | 3.24 (2.00, 5.23) |
|  | Bone mets vs mets -ve (Rectum + LT Colon) | 1257 | 133 | 1124 | 9.48 | 3.31E-10 | 1647 | 146 | 1501 | 8.47 | 3.42E-09 | | 4.10 (2.49, 6.77) |
| rs16889360 | Bone mets vs non-bone mets (Rectum + LT Colon) | 1034 | 133 | 901 | 6.58 | 2.66E-07 | 1295 | 146 | 1149 | 7.52 | 3.00E-08 | | 3.66 (2.20, 6.09) |
|  | Bone mets vs mets -ve (Rectum + LT Colon) | 1257 | 133 | 1124 | 9.46 | 3.51E-10 | 1647 | 146 | 1501 | 8.03 | 9.40E-09 | | 4.50 (2.72, 7.46) |
|  | Bone mets vs mets -ve (overall) | 1545 | 150 | 1395 | 7.97 | 1.07E-08 | 2057 | 171 | 1886 | 6.47 | 3.37E-07 | | 3.24 (2.00, 5.23) |
| rs4365724 | Bone mets vs mets -ve (Rectum + LT Colon) | 1257 | 133 | 1124 | 7.26 | 5.50E-08 | 1647 | 146 | 1501 | 4.65 | 2.26E-05 | | 2.78 (1.70, 4.54) |
| rs2919408 | Bone mets vs mets -ve (Rectum + LT Colon) | 1257 | 133 | 1124 | 7.26 | 5.50E-08 | 1647 | 146 | 1501 | 4.65 | 2.26E-05 | | 2.78 (1.70, 4.54) |
| rs1491041 | Bone mets vs mets -ve (Rectum + LT Colon) | 1257 | 133 | 1124 | 6.59 | 2.60E-07 | 1647 | 146 | 1501 | 7.43 | 3.69E-08 | | 1.99 (1.35, 2.93) |
| rs994544 | I/II | 1089 | 251 | 838 | 6.38 (Dom) | 4.20E-07 | 1421 | 314 | 1107 | 5.52 | 2.99E-06 | | 3.02 (1.86, 4.89) |
| rs7808582 | Male only, stage III | 593 | 302 | 291 | 7.21 | 6.16E-08 | 785 | 380 | 405 | 4.83 | 1.49E-05 | | 1.97 (1.44, 2.69) |
| rs4953913 | Peritoneal mets vs non-peritoneal mets (Female,Rectum + LT Colon) | 426 | 94 | 332 | 7.37 | 4.31E-08 | 526 | 114 | 412 | 5.69 | 2.03E-06 | | 2.45 (1.56, 3.87) |
| rs6564734 | Lung +/@ Liver mets vs no mets, stage III | 995 | 442 | 553 | 6.55 | 2.81E-07 | 1300 | 529 | 771 | 4.96 | 1.09E-05 | | 1.75 (1.32, 2.31) |
