## Supplemental Table S4 for "Functional annotation with expression validation identifies novel metastasis-relevant genes from post-GWAS risk loci in sporadic colorectal carcinomas"

**Table S4**: **Sample sizes of overall and disease subtype datasets**

| S/N | Disease Subtype | Sample Size | | |
| --- | --- | --- | --- | --- |
|  |  | Total | Met+ve | Met-ve |
| 1. | Overall | 2677 | 1282 | 1395 |
| 2. | Left-sided | 2147 | 1027 | 1120 |
| 3. | Stage I/II | 1089 | 251 | 838 |
| 4. | Left-sided stage I/II | 890 | 210 | 680 |
| 5. | Stage III | 1086 | 535 | 551 |
| 6. | Left-sided stage III | 861 | 427 | 434 |
| 7 | Left-sided liver and/or lung stage I/II/III | 1650 | 537 | 1113 |
| 8 | Bone metastasis | 1545 | 150 | 1395 |
| 9 | Left-sided peritoneal-metastasis | 1326 | 206 | 1120 |
| 10 | Male | 1476 | 729 | 747 |
| 11 | Female | 978 | 331 | 647 |
